# Cleft lip sidedness and the association with additional congenital malformations

**DOI:** 10.1101/2024.02.14.24302817

**Authors:** Matthew Fell, Kate J Fitzsimons, Mark J Hamilton, Jibby Medina, Sophie Butterworth, Min Hae Park, Jan Van der Meulen, Sarah Lewis, David Chong, Craig JH Russell

**Author notes:** Joint First Authors (Matthew Fell, Kate J Fitzsimons). These authors contributed equally to this work. Corresponding author: Matthew Fell, The Cleft Collective, Bristol Dental School, University of Bristol, Oakfield House, Oakfield Grove, Bristol, BS8 2BN, United Kingdom.

## Abstract

**Objective:** To investigate the association between the sidedness of orofacial clefts and additional congenital malformations.

**Design:** Linkage of a national registry of cleft births to national administrative data of hospital admissions

**Setting:** National Health Service, England

**Participants:** 2,007 children born with cleft lip +-alveolus (CL+-A) and 2,724 with cleft lip and palate (CLP) born between 2000 and 2012.

**Main outcome measure:** The proportion of children with ICD-10 codes for additional congenital malformations by the sidedness (left, right or bilateral) of orofacial clefts.

**Results:** For CL+-A phenotypes, there was no evidence for a difference in the prevalence of additional anomalies between left (22%, reference), right (22%, aOR 1.02, 95% CI 0.80 to 1.28; p= 0.90) and bilateral clefts (23%, aOR 1.09, 95% CI 0.75 to 1.57; p= 0.66). For CLP phenotypes, there was evidence of a lower prevalence of additional malformations in left (23%, reference) compared to right (32%, aOR 1.54, 95% CI 1.25 to 1.91; p <0.001) and bilateral clefts (33%, aOR 1.64, 95% CI 1.35 to 1.99; p<0.001).

**Conclusions:** The prevalence of additional congenital malformations was similar across sidedness subtypes with CL+-A phenotypes but was different for sidedness subtypes within CLP cases. These data support the hypothesis that CL+-A has a different underlying aetiology from CLP and that within the CLP phenotype, right sided CLP may lie closer in aetiology to bilateral CLP than it does to left sided CLP.

## Background

There are consistent trends for sidedness in clefts of the lip with or without palate (CL+-P) that has been termed ‘directional asymmetry’.^1,2^ The prevalence of left unilateral CL+-P is twice that of right unilateral CL+-P.^2,3^ Bilateral CL+-P occurs less commonly than unilateral CL+-P, with a quoted prevalence ratio of 6:3:1 for left/right/bilateral sidedness, respectively.^4^ Directional asymmetry is not a unique phenomenon to CL+-P, as left sided predominance is also observed in preauricular skin tags, congenital hip dysplasia and absent upper limb, whereas right sided predominance is seen in microtia and pre-auricular sinus.^5^ Both genetic and environmental factors have been proposed in the aetiology of sidedness in CL+-P, yet mechanisms are currently poorly understood.^6^

Orofacial clefts can occur in isolation or alongside additional congenital malformations (ACMs). Congenital malformations have been defined as defects involving a functional, structural, morphological or positional anomaly of a whole or part of a body organ system that tends to be macroscopic and occurs before birth.^7^ ACMs can occur as part of a syndrome, which may arise due to a change within the genetic material (including single-gene disorders or larger chromosome anomalies) and/or teratogenic exposures.^8^ The study and identification of co-occurring ACMs is illuminating because it can give insights into the underlying aetiology of orofacial clefts,^9^ and in turn, facilitates the design of bespoke treatment pathways.^10^

The United Kingdom (UK) is well positioned to investigate the association of CL+-P sidedness with ACMs due to the systematic collection of data on cleft at a nationwide level within a centralised healthcare system, which is free at the point of access. We have previously published the range and frequency of ACMs in 9,403 children born with orofacial clefts and noted a difference in prevalence of ACMs between cleft lips +-alveolus (CL+-A) and cleft lip and palate (CLP).^11^ This study aims to investigate whether there is any difference in the prevalence of ACMs by sidedness in both CL+-A and CLP using two national datasets in England, linked at the individual patient level.

## Methods

### Data source

The study cohort was identified in the Cleft Registry and Audit NEtwork (CRANE) Database which is a national registry of all live-born children with an orofacial cleft (OFC) in the UK. Cases registered are most commonly identified antenatally or at birth (84%) and overall case ascertainment has recently been calculated as above 95%.^12^ Children whose parents had given consent for their child’s records to be linked to other datasets were eligible to be linked to the Hospital Episode Statistics (HES) database.

The HES database^13^ contains records on all diagnoses and treatments made and given during admissions to National Health Service (NHS) hospitals in England. HES is derived from routine submission of data from each individual NHS hospital admission, primarily for the purposes of payment for and commissioning of healthcare in England.^14^ Data is inputted locally by professional health coders before undergoing central data quality processing for validity and data cleaning checks prior to being published. The linked dataset contained records on births from January 1^st^ 2000 up to December 31^st^ 2012 and hospital admissions up to March 31^st^ 2015, to allow sufficient time for the existence of ACMs to be identified and recorded.

### Cleft subtype classification

Children born with either a unilateral or bilateral cleft lip with or without a cleft palate were included. In CRANE, cleft phenotypes and sidedness are categorised according to the LAHSAL classification.^15^ In HES, cleft type was assigned according to the presence of selected procedure codes (Classification of Interventions and Procedures, OPCS 4.5) and/or diagnosis codes (tenth revision of the International Classification of Diseases, ICD-10) ^16^ in any of the available HES records. A stepwise, hierarchical approach was employed. First, the cleft reconstruction procedure codes (F03, F29, F32) were used to identify cleft type groups: CL+-A, CLP and cleft palate only. Second, the diagnosis code was used to distinguish between unilateral and bilateral cases. Children with cleft palate only and those whose cleft type was discordant between the two data sources were excluded from the study cohort.

### Additional congenital malformations

Congenital malformations were classified based on the International Classification of Disease Version 10 (ICD-10) codes ^16^ which are reported in HES data. ICD-10 is considered the international standard diagnostic classification for disease and is developed and maintained by the World Health Organisation. Congenital malformations, deformations and chromosome abnormalities are coded Q00-Q99 and are categorised according to the body or organ system they affect (see Appendix 1).

### Data Analysis

The proportion of children with ICD-10 codes for congenital malformations (listed in Appendix 1) was examined. Differences in the percentage of children with ACMs between sidedness subgroups were initially explored using the Pearson chi-square test of independence. Children born with CL+-A were analysed separately from children born with CLP, as previous findings indicated different proportions of ACMs between the two phenotypes,^11^ and in line with developing knowledge to suggest that these cleft subtypes should be considered as distinct aetiological entities.^17–20^

The number of different body or organ systems with malformations was summed for each child, and the percentage of children with ACMs affecting two or more body systems was reported by cleft subtype. ‘Chromosomal abnormalities not elsewhere specified’ (Q90-Q99) were not included in these particular analyses, as the aim was to identify specific body systems affected by physical malformations, rather than the underlying cause. Similarly, while certain chromosomal diagnoses may be associated with high likelihood of particular malformations, it is not a given that the associated anomalies will be present, so we did not assume physical anomalies existed based on the presence of chromosomal anomalies.

The association between cleft sidedness (left, right or bilateral) and the presence of ACMs was analysed using logistic regression to estimate effect size, using left sided clefts as a reference group and adjusting for patient sex. Additional adjustment was made by adding ethnicity into the regression model, but due to incomplete ethnicity data reducing the sample size, these results are reported as supplementary data only. Odds ratios, confidence intervals and p values are reported and interpreted as continuous measures of the strength of evidence against the null hypothesis.^21^ Statistical analysis was performed in Stata nV.17 (Statacorp).

### Ethical Considerations

This study is exempt from NHS Research Authority ethics approval as it involves the analysis of an existing anonymised dataset that is collected for the purpose of service evaluation.^22^

## Results

### Patient Characteristics

5,940 CRANE-registered children were born alive in England with a CL+-P between 1^st^ January 2000 and 31^st^ December 2012. 5,640 (95%) CRANE records were successfully linked to HES records but 1,209 were excluded due to cleft phenotype discordance between the two data sources. The final cohort in this study included 4,731 children born with CL+-P and of these, 42% had CL+-A and 58% had CLP (see Table 1). Among children with unilateral clefts, the ratio between left and right-sides was similar between those with CL+-A and those with CLP (65%:35% vs 63%:37% respectively, p=0.279), However, the distribution of left, right and bilateral clefts differed between the CL+-A and CLP groups (59%, 32%, 9% vs. 43%, 25%, 32% respectively, p<0.001).

**Table 1.**
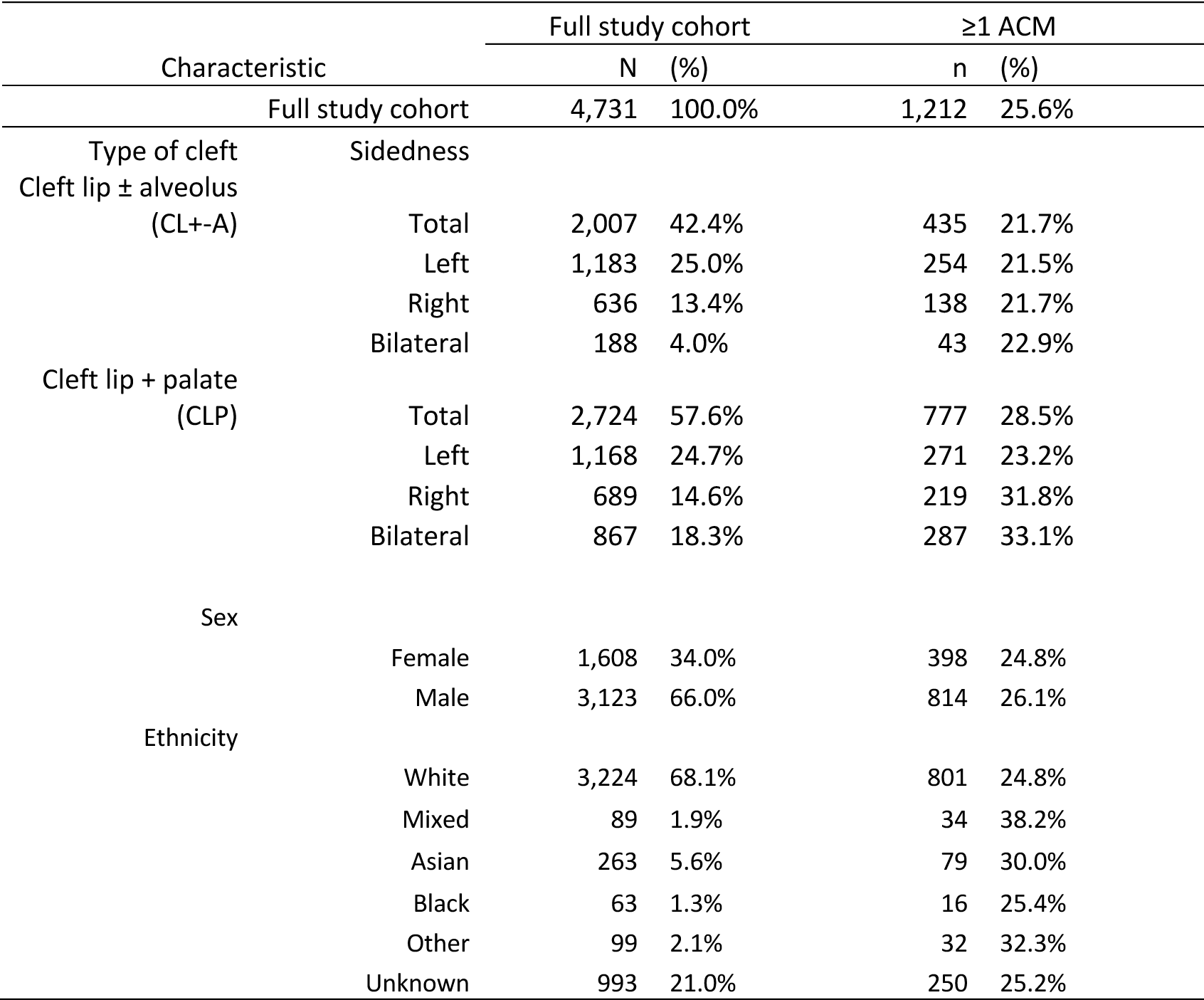
Characteristics of the children included in the analyses and the number and percentage of those with ≥1 additional congenital malformations (ACM)

66% of the cohort was male. This proportion differed between CL+-A and CLP groups (63% vs 68%, p<0.001) but there was no difference in the proportion of ACMs between males and females (26% vs 25% p=0.33). Among those with reported ethnicity (79%), the proportion identified as White British was similar between the CL+-A and CLP groups (87% vs 86%, p=0.58). The overall proportion of ACMs was lowest in White British and Black ethnicities and highest in Mixed, Asian and ‘Other’ ethnicity groups (see Table 1).

### Cleft lip +-alveolus (CL+-A)

When compared to the proportion of ACMs in left sided CL+-A (22%), there was no difference in the proportion of ACMs in either right UCL+-A (22%, aOR 1.02, 95% CI 0.80 to 1.28; p=0.90) or bilateral UCL+-A (23%, aOR 1.09, 95% CI 0.75 to 1.57; p=0.66) as seen in Table 2. Additional adjustment for ethnicity did not alter this finding (see Supplementary Table 1).

**Table 2.**
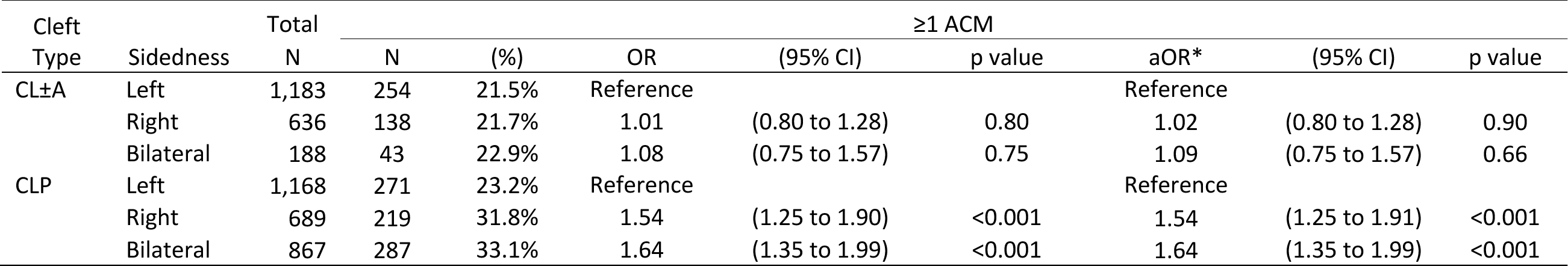
The association of additional congenital malformations, according to cleft type (Cleft lip ± alveolus (CL±A) and Cleft Lip and Palate (CLP)) and sidedness. Effect estimate is the odds ratio (OR) with 95% confidence intervals (Cis) and p values. *Adjusted for sex.

| Cleft Type | Sidedness | Total N | ≥1 ACM |  |  |  |  |  |  |  |
| --- | --- | --- | --- | --- | --- | --- | --- | --- | --- | --- |
|  |  |  | N | (%) | OR | (95% CI) | p value | aOR* | (95% CI) | p value |
| CL±A | Left | 1,183 | 254 | 21.5% | Reference |  |  | Reference |  |  |
|  | Right | 636 | 138 | 21.7% | 1.01 | (0.80 to 1.28) | 0.80 | 1.02 | (0.80 to 1.28) | 0.90 |
|  | Bilateral | 188 | 43 | 22.9% | 1.08 | (0.75 to 1.57) | 0.75 | 1.09 | (0.75 to 1.57) | 0.66 |
| CLP | Left | 1,168 | 271 | 23.2% | Reference |  |  | Reference |  |  |
|  | Right | 689 | 219 | 31.8% | 1.54 | (1.25 to 1.90) | <0.001 | 1.54 | (1.25 to 1.91) | <0.001 |
|  | Bilateral | 867 | 287 | 33.1% | 1.64 | (1.35 to 1.99) | <0.001 | 1.64 | (1.35 to 1.99) | <0.001 |

When ACMs were stratified by the ICD-10 body system subgroups, the ten subcategories were present in similar proportions among children with a left, right or bilateral CL+-A in many of the body systems (See Table 3). When compared to the left CL+-A reference group, respiratory anomalies were more common in children with a right CL+-A (OR 1.54, 95% CI 1.01 to 2.34; P=0.04), whereas the bilateral CL+-A phenotype had a higher proportion of eye, ear, face anomalies (OR 2.78, 95% CI 1.25 to 6.17; P=0.01) and circulatory anomalies (OR 2.73, 95% CI 1.49 to 4.99; P=0.001), although the evidence for these associations was weak given that we performed multiple tests.

**Table 3.**
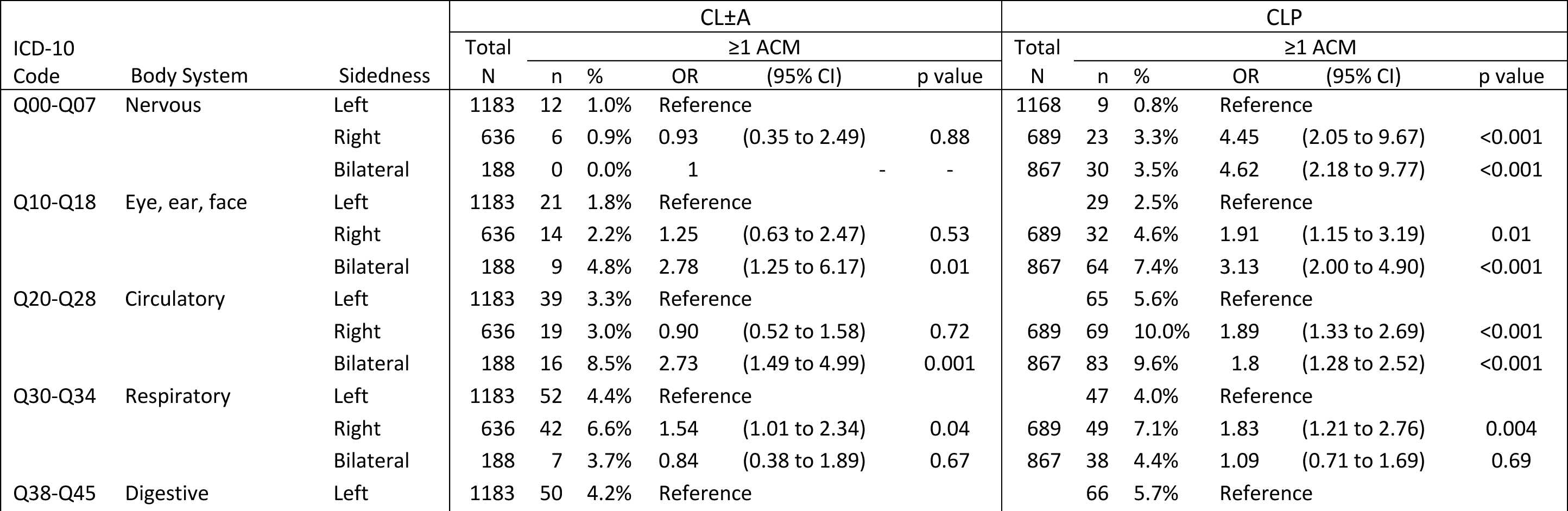
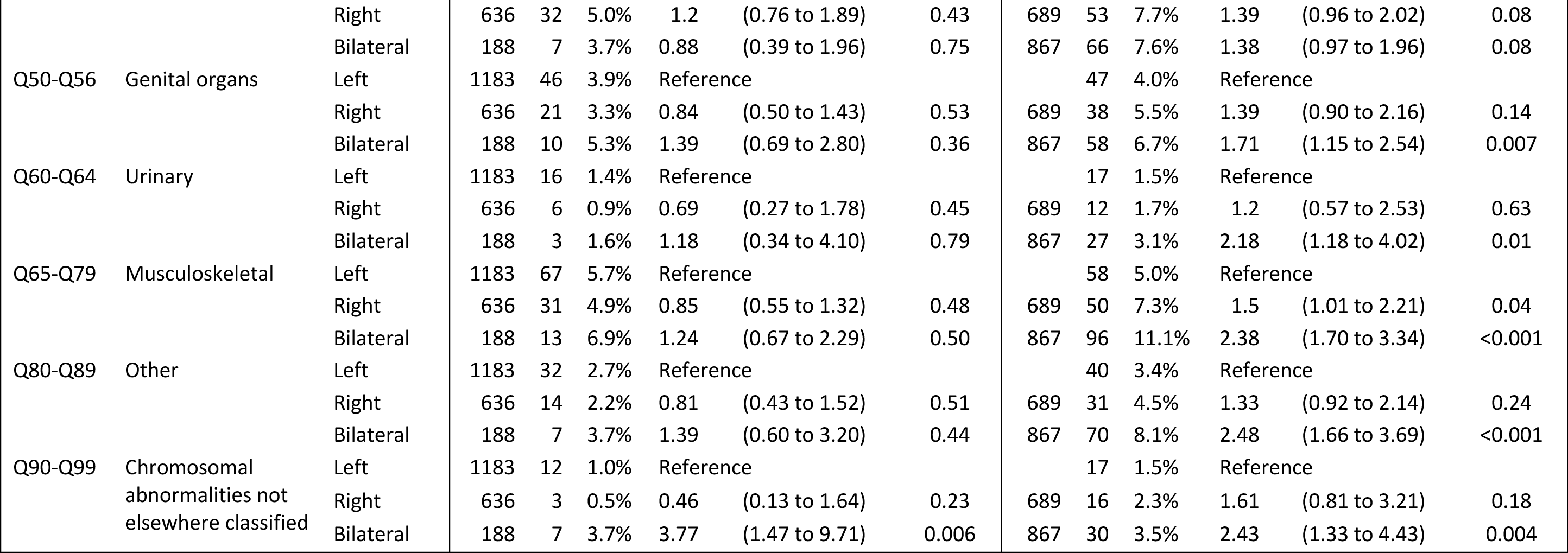
The association of additional congenital malformations according to ICD-10 body system classification, by cleft type (cleft lip ± alveolus (CL±A) and cleft lip and palate (CLP)) and sidedness. Effect estimate is the odds ratio (OR) with 95% confidence intervals (Cis) and p values

### Cleft lip and palate (CLP)

When compared to the proportion of ACMs in left sided CLP (23%) there was a higher proportion of ACMs amongst both right CLP (32%, aOR 1.54, 95% CI 1.25 to 1.91; p<0.001) and bilateral CLP (33%, aOR 1.64, 95% CI 1.35 to 1.99; p<0.001) as seen in Table 2. Additional adjustment for ethnicity did not alter this trend (Supplementary Table 1).

When ACMs were stratified by the ICD-10 body system subgroups, each of the ten subcategories of ACMs were more prevalent in both right and bilateral CLP when compared to the left CLP reference (see Table 3) The direction of the effect estimate for right and bilateral clefts was consistent for each body system, but the strength of association for ACMs compared to left CLP varied depending on the body system.

### Number of systems affected by additional congenital malformations

Table 4 shows the number and percentage of children who had multiple (≥2) body systems (e.g. nervous, eye/ear/face, circulatory, respiratory, digestive, genital, urinary and musculoskeletal systems) affected by ACMs. Among those with CL+-A, the proportion of children who had malformations across multiple body systems when compared to left CL+-A (5%) was similar for right CL+-A (4%, aOR 0.74, 95% CI 0.44 to 1.22; p=0.23) but was greater for bilateral CL+-A (10%, aOR 2.17, 95% CI 1.25 to 3.79; p<0.001). Among those with CLP, the proportion of children who had ACMs across multiple body systems when compared to left CLP (6%), was greater for both right CLP (11%, aOR 1.76, 95% CI 1.25 to 2.46; p<0.001) and bilateral CLP (14%, aOR 2.45, 95% CI 1.81 to 3.31; p<0.01).

**Table 4.**
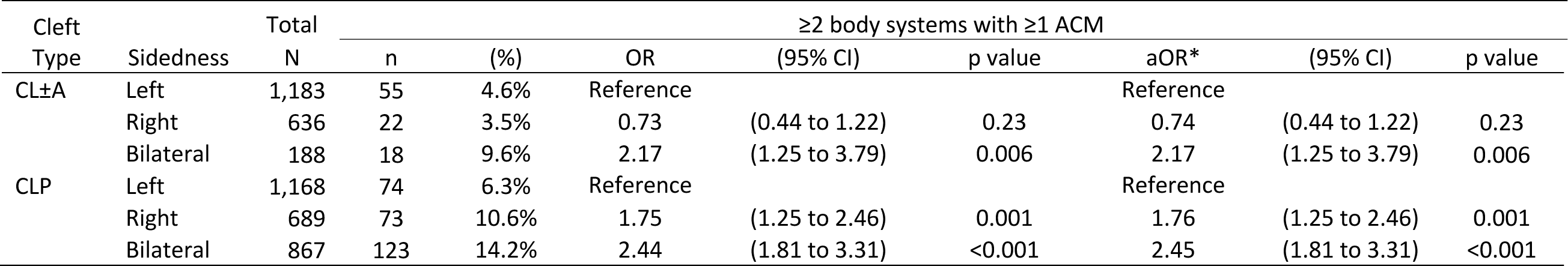
Number of body systems affected by additional congenital malformations, according to cleft type (Cleft lip ± alveolus (CL±A) and Cleft <u>lip and palate (CLP)) and sidedness. Effect estimate is the odds ratio (OR) with 95% confidence intervals (Cis) and p values. *Adju</u>sted for sex.

## Discussion

### Summary of findings

In this national sample of 4,731 children born with CL+-P, we were able to examine the association of cleft sidedness with the prevalence of additional congenital malformations as coded by the ICD-10 classification. The prevalence of ACMs for children born with CL+-A ranged between 22% to 23% and whilst there was no evidence to suggest differences between left, right or bilateral sidedness, there was evidence to suggest a greater proportion of ACMs in multiple body systems in bilateral CL+-A (10%) compared to left (5%) or right (4%) CL+-A.

The prevalence of ACMs for children born with CLP was different according to sidedness, with 23% of left sided CLP having at least one additional malformation compared to 32% in right CLP and 33% in bilateral CLP. The subcategorization of ICD-10 malformation codes into body systems within the CLP subtype, consistently showed a higher prevalence of malformations for right and bilateral CLP compared to left CLP in each body system, reducing the likelihood that this trend is a chance finding. In addition, the proportion of ACMs in multiple body systems was greater for both right (11%) and bilateral (14%) CLP compared to left CLP (6%).

### Comparison to previous studies

Pereira et al.,^10^ analysed the presence of additional congenital malformations in 701 children with orofacial clefts from a Portuguese database, 343 of whom had unilateral CL+-P. Among children with additional malformations, there was a higher prevalence of right CL+-P compared to the group without malformations (13% vs 11%; P<0.001). The authors did not stratify individual malformations by sidedness but, for the entire cohort, head and neck anomalies were found to be the most common, followed by cardiovascular malformations.

Gallagher et al.,^23^ reported a greater prevalence of syndromes amongst 155 children born with a right sided CL+-P compared to 276 born with a left sided CL+-P (OR 1.9, 95% CI 0.7 to 4.7). The imprecise estimate may have been linked to the small sample size and subjective method of identifying syndromes. The authors concluded that children born with right sided CL+-P were more likely to be born with ACMs and whilst this is consistent with our data, they did not subcategorise the clefts into CL+-A and CLP subtypes.

### Interpretation of the data

Our data can be used to consider the current theories that have been suggested for the aetiology of sidedness in CL+-P. Various case control and genome wide association studies (GWAS) have highlighted distinct genetic variations associated with left versus right sided CL+-P over the past two decades.^20,24–28^ For example, using a GWAS approach, Curtis et al.,^20^ found evidence that a polymorphism at locus 4q28, close to the *FAT4* gene, influenced sidedness in unilateral CL+-A, whilst no significant effect was detected for this locus in unilateral CLP. For decades, since pioneering work by Fogh-Anderson,^29^ it has been recognised that the aetiology of cleft palate only is likely different to that of CL+-P. Further, subsequent evidence has emerged to suggest distinct aetiologies between CL+-A and CLP.^17^ Our data contributes further evidence that sidedness may also be distinct between CL+-A and CLP subtypes.

The bilateral CL+-P cohort in this study provides a useful comparative group for the unilateral Cl+-P cohort. First, cleft lip sidedness ratios are often quoted in the literature as 6:3:1 for left:right:bilateral^4^ and whilst this was true in our CL+-A cohort, the proportion of bilateral CLP was similar to right CLP, as has been previously reported in other orofacial cleft epidemiology studies.^2,30^ Second, for the CL+-A subtype, the prevalence of ACMs is similar irrespective of left, right or bilateral sidedness, whereas for the CLP subtype the prevalence of ACMS is higher for right and bilateral CLP compared to left CLP.

The amalgamation of all cleft subtypes included in our main and supplementary analysis can be presented as a spectrum using left CL+-A as a reference (see Figure 1 and supplementary Table 2) and demonstrates right and bilateral CLP standing apart from the other cleft lip subtypes, according to the proportion of associated ACMs. In two studies investigating the co-occurrence of congenital dental anomalies with cleft sidedness, there was a reported higher prevalence of left maxillary lateral incisor agenesis in right CL/P compared with the reverse scenario.^31,32^ Both authors suggested the occurrence of a missing lateral incisor on the non-cleft side may represent a bilateral genotype that did not fully manifest and that right sided CL+-P may be more likely to correspond to a milder phenotype of bilateral clefts.^33,34^

**Figure 1.**
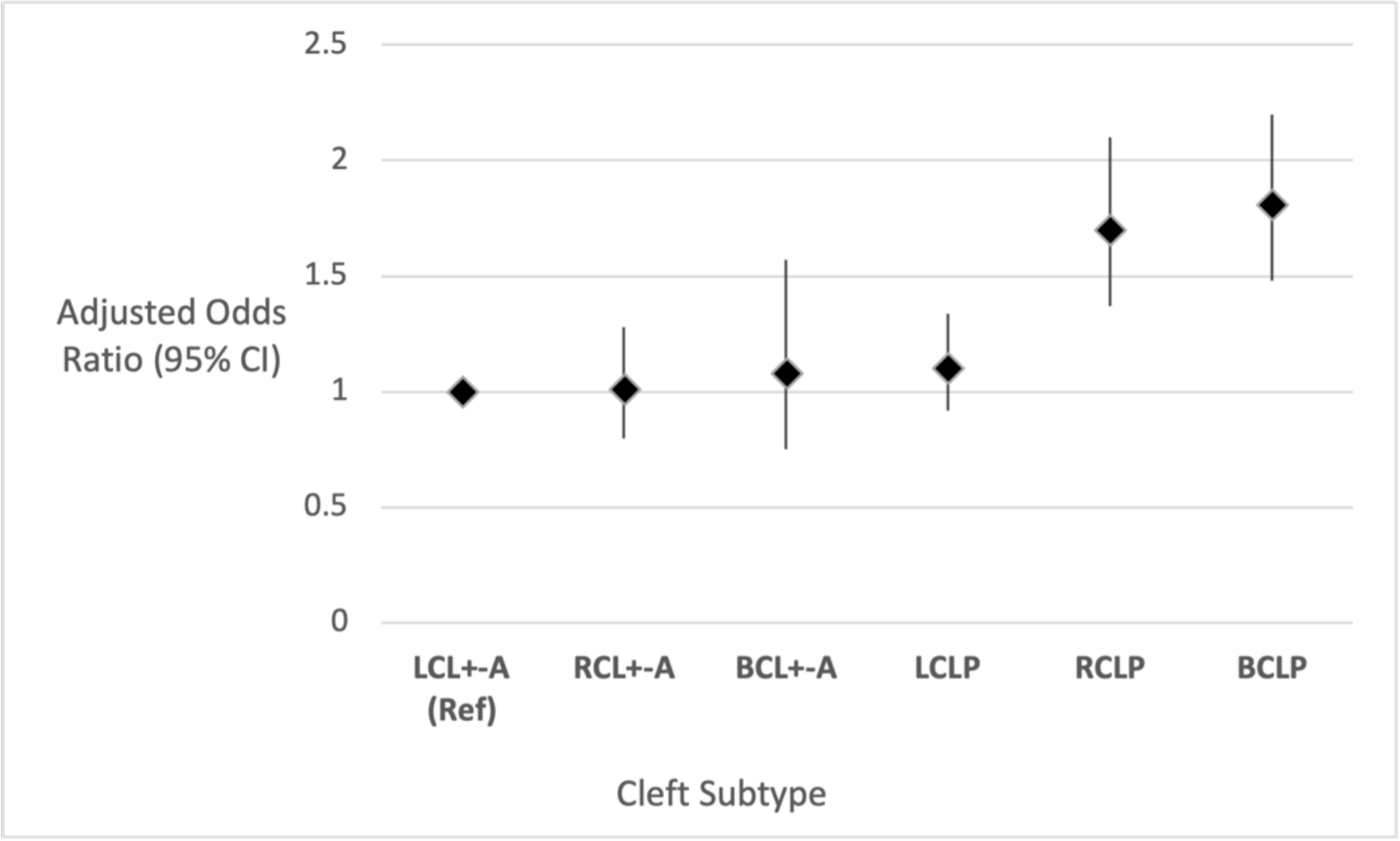
The association of additional congenital malformations across the spectrum of unilateral and bilateral cleft lip with or without cleft palate. Groups are categorised according to sidedness left (L), right (R) and bilateral (B) and the cleft subtypes cleft lip +-alveolus (CL+-A) and cleft lip and palate (CLP). Left unilateral cleft lip +-A (LUCL+-A) is used as the reference group. Effect estimates displayed as Odds Ratio (OR) with 95% Confidence Intervals (CI), adjusted for sex.

Investigations into the aetiology of orofacial cleft have reached a broad consensus that non-syndromic clefts arise within a multifactorial background.^33,34^ Right sided CLP and bilateral CLP occur less commonly than left sided CLP, and may represent more severe phenotypes, requiring a higher threshold of genetic and environmental influences for them to occur.

### Strengths and Limitations

This is a large cohort of standardised national data within a centralised and nationalised health care system, and therefore represents a unique resource. The CRANE registry has high case ascertainment,^12^ and given its national coverage is likely to be representative of children born with an orofacial in England during the study period. A minimum follow-up of 2.25 years in HES data allowed sufficient time for congenital anomalies to be identified after birth. Discordance in cleft subtypes in one fifth of the patient records between the datasets was a weakness as it resulted in the exclusion of otherwise successfully linked data.

The ICD-10 categorisation system of malformations is advantageous because it is an international system that is widely accessible and facilitates thorough documentation of anatomical detail. However, despite the ICD-10 being regarded as the international standard, a variety of different classifications have been used in the literature, which makes comparison difficult ^10,35^. The ICD-10 anatomical subcategorization is not necessarily clinically intuitive. Whilst the use of ICD-10 codes allowed us to report many congenital malformations, HES restricts the entry of these codes to 4 characters (e.g. Q87.0), which means that some codes were not sensitive enough to distinguish between certain diagnoses (for example, Pierre Robin Sequence and Goldenhar syndrome share the same 4-character ICD-10 code). Furthermore, ICD-10 codes utilised in HES tend to focus on a physical diagnosis or phenotype, rather than the underlying genetic cause. This means the prevalence of specific genetic and/or syndromic diagnoses associated with orofacial clefts and other congenital malformations could not be reported.

This study was restricted to children born alive. It was not possible to include spontaneous abortions, elective terminations and stillborn foetuses. Furthermore, as there is no standard protocol for evaluating other body systems for anomalies in children presenting with a cleft in England, there may well be subclinical and untreated anomalies that have been missed in the study population. Whilst there is no reason to suggest that the prevalence of missed ACMs would differ according to cleft sidedness, the true prevalence of additional malformations is likely to be underrepresented.

Finally, we adjusted for the potential confounders of sex and ethnicity as these variables were readily available in the data set. Adjustment for these confounders did not alter the trends observed indicating that the reported findings were not explained by these characteristics. The proportion of children with additional malformations according to ethnic background was limited by missing data and slightly lower representation of minority ethnic groups in this cohort as compared to last UK census.^36^ Furthermore, we were not able to add additional confounding factors to our model, which could be important in cleft aetiology, such as maternal age, Body Mass Index, and folic acid consumption.

### Further work

Our study demonstrates the importance of any study on cleft sidedness being explicit about the inclusion or exclusion of children born with ACMs, as the proportion of ACMs will likely be different for children born with left, right and bilateral CLP. Further work to understand the aetiology of sidedness in CL+-P should stratify participants by CL+-A and CLP subtypes. Furthermore, future studies should make efforts to distinguish individuals within cleft cohorts who have a known monogenic or chromosomal diagnosis, from those whose cleft is assumed to have a complex or multifactorial cause, since genetic influences may be overlapping or distinct between these groups.

Our results suggest a research focus on left CLP versus right and bilateral CLP is warranted. Work to investigate genetic factors should be expanded to include large scale GWAS studies. DNA methylation has been shown to play a critical role in establishing laterality of the body plan during early embryogenesis in zebrafish models,^37^ and methylation patterns have been shown to differ by CL+-A, CLP and CP subtype in humans,^17^ yet we are not aware of its use in orofacial cleft sidedness research and studying methylation patterns could offer important insights in this area. This may help to enhance our understanding of how cleft sub-phenotypes are classified and provide more information to support or refute the suggestion of right sided CLP and bilateral CLP being different phenotypic expressions of the same condition.

### Conclusion

This large population-based study provides evidence for an increased risk of additional congenital malformations among right and bilateral CLP compared to left CLP. This was not the case for CL+-A, where additional congenital malformations were equivalent according to cleft sidedness. This supports a growing evidence base to suggest CLP has a different aetiology when compared with CL+-A and that these subtypes of cleft should not be considered as a single entity. Furthermore, this data suggests that right CLP may lie closer in aetiology to bilateral CLP than it does to left sided CLP. This finding may help to explain why right sided orofacial clefts are less prevalent than left sided orofacial clefts, although further work is required to explain, define and understand this.

## Data Availability

All data produced in the present work are contained in the manuscript

---

**Appendix 1:**
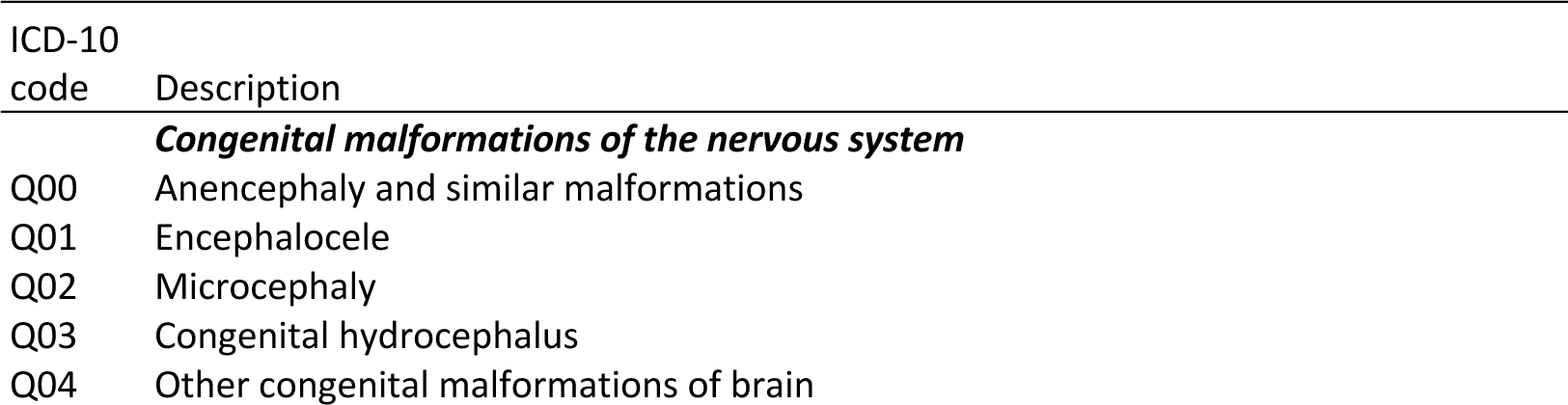
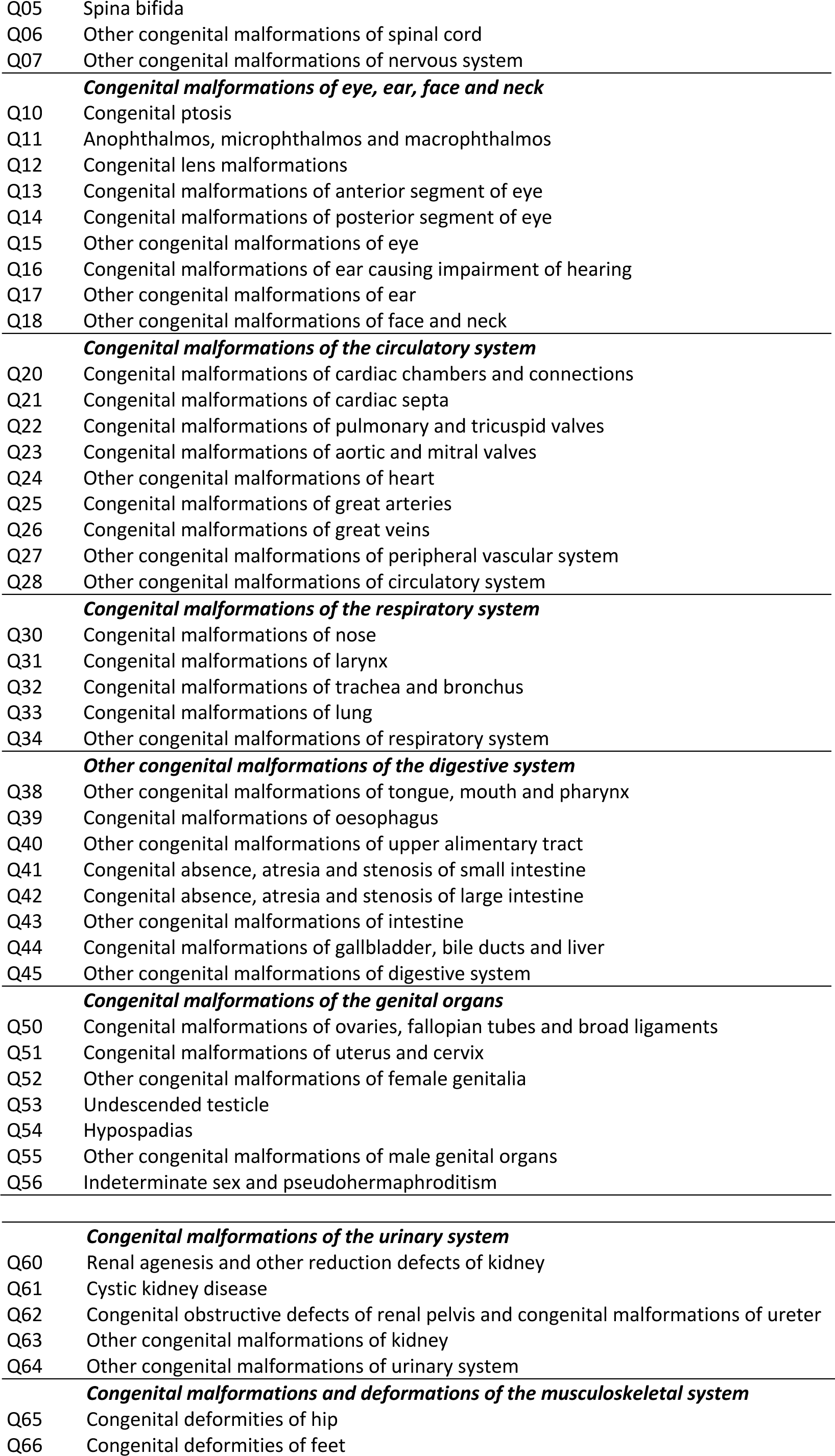
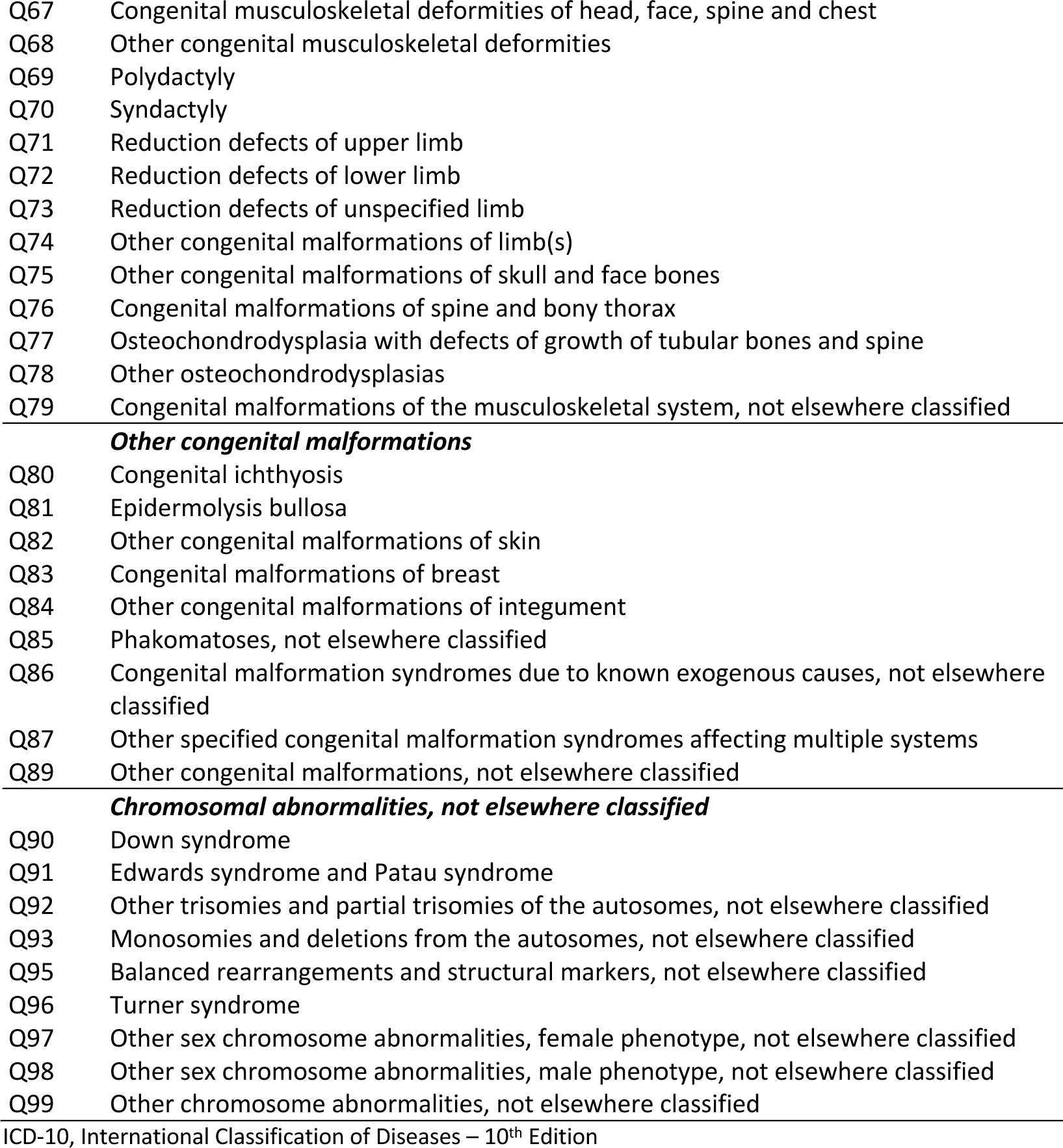
Diagnostic codes used to identify congenital malformations, and the number (%) of children in the cleft cohort with these codes in their Hospital Episode Statistics history.

## Supplementary tables

**Supplementary Table 1.** Odds ratios (95% confidence intervals) for additional congenital malformations, according to cleft type and sidedness. Adjustment made for sex (male vs female) and ethnicity (White British vs BME). Reduced total numbers due to missing ethnicity data.

| Cleft type | Sidedness | Total | ≥1 ACM |  |  |  |  |  |  |  |
| --- | --- | --- | --- | --- | --- | --- | --- | --- | --- | --- |
|  |  | N | n | % | OR | (95% CI) | p value | aOR* | (95% CI) | p value |
| CL±A | Left | 922 | 192 | 20.8% | Reference |  |  | Reference |  |  |
|  | Right | 504 | 111 | 22.0% | 1.07 | (0.82 to 1.40) | 0.60 | 1.01 | (0.80 to 1.28) | 0.91 |
|  | Bilateral | 150 | 36 | 24.0% | 1.2 | (0.80 to 1.80) | 0.38 | 1.07 | (0.74 to 1.54) | 0.73 |
| CLP | Left | 934 | 217 | 23.2% | Reference |  |  | Reference |  |  |
|  | Right | 553 | 177 | 32.0% | 1.56 | (1.23 to 1.97) | <0.001 | 1.57 | (1.24 to 1.99) | <0.001 |
|  | Bilateral | 675 | 229 | 33.9% | 1.7 | (1.36 to 2.11) | <0.001 | 1.72 | (1.38 to 2.14) | <0.001 |

\*Adjusted for sex and ethnicity

**Supplementary Table 2.** Proportion of additional congenital malformations across of the spectrum of unilateral and bilateral cleft lip with or without cleft palate using left UCL±A as the reference. Adjusted OR and confidence intervals used for Figure 1.

| Cleft type & sidedness | Total | ≥1 ACM |  |  |  |  |  |  |  |
| --- | --- | --- | --- | --- | --- | --- | --- | --- | --- |
|  | N | N | (%) | OR | (95% CI) | p value | aOR* | (95% CI) | p value |
| LCL±A | 1,183 | 254 | 21.5% | Reference |  |  | Reference |  |  |
| RCL±A | 636 | 138 | 21.7% | 1.01 | (0.80 to 1.28) | 0.91 | 1.01 | (0.80 to 1.28) | 0.91 |
| BCL±A | 188 | 43 | 22.9% | 1.08 | (0.75 to 1.57) | 0.66 | 1.08 | (0.75 to 1.57) | 0.66 |
| LUCLP | 1,168 | 271 | 23.2% | 1.10 | (0.91 to 1.34) | 0.31 | 1.10 | (0.91 to 1.34) | 0.32 |
| RUCLP | 689 | 219 | 31.8% | 1.70 | (1.38 to 2.11) | <0.001 | 1.70 | (1.37 to 2.10) | <0.001 |
| BCLP | 867 | 287 | 33.1% | 1.81 | (1.48 to 2.21) | <0.001 | 1.81 | (1.48 to 2.20) | <0.001 |

\*Adjusted for sex

## References

1. Nagase Y, Natsume N, Kato T, Hayakawa T. Epidemiological Analysis of Cleft Lip and/or Palate by Cleft Pattern. J Maxillofac Oral Surg. 2010;9(4):389–395. doi:10.1007/s12663-010-0132-6

2. Mossey PA, Modell B. Epidemiology of Oral Clefts 2012: An International Perspective. In:; 2012:1–18. doi:10.1159/000337464

3. Mossey PA, Little J, Munger RG, Dixon MJ, Shaw WC. Cleft lip and palate. The Lancet. 2009;374(9703):1773-1785. doi:10.1016/S0140-6736(09)60695-4

4. Martin S V., Swan MC. An essential overview of orofacial clefting. Br Dent J. 2023;234(12):937–942. doi:10.1038/s41415-023-6000-9

5. Paulozzi LJ, Lary JM. Laterality patterns in infants with external birth defects. Teratology. 1999;60(5):265–271. doi:10.1002/(SICI)1096-9926(199911)60:5<265::AID-TERA7>3.0.CO;2-H

6. Fell M, Bradley D, Chadha A, et al. Sidedness in Unilateral Orofacial Clefts: A Systematic Scoping Review. The Cleft Palate Craniofacial Journal. Published online December 13, 2023. doi:10.1177/10556656231221027

7. Impellizzeri A, Giannantoni I, Polimeni A, Barbato E, Galluccio G. Epidemiological characteristic of Orofacial clefts and its associated congenital anomalies: Retrospective study. BMC Oral Health. 2019;19(1). doi:10.1186/s12903-019-0980-5

8. Vallino-Napoli LD, Riley MM, Halliday JL. An Epidemiologic Study of Orofacial Clefts With Other Birth Defects in Victoria, Australia.

9. Sárközi A, Wyszynski DF, Czeizel AE. Oral clefts with associated anomalies: Findings in the Hungarian Congenital Abnormality Registry. BMC Oral Health. 2005;5. doi:10.1186/1472-6831-5-4

10. Pereira A V., Fradinho N, Carmo S, et al. Associated malformations in children with orofacial clefts in Portugal: A 31-year study. Plast Reconstr Surg Glob Open. 2018;6(2). doi:10.1097/GOX.0000000000001635

11. Fitzsimons KJ, Hamilton MJ, van der Meulen J, et al. Range and Frequency of Congenital Malformations Among Children With Cleft Lip and/or Palate. Cleft Palate-Craniofacial Journal. Published online 2022. doi:10.1177/10556656221089160

12. Cleft Registry and Audit NEtwork. 2023 Annual Report. Accessed 14th February 2024. https://www.crane-database.org.uk/content/uploads/2024/01/CRANE-2023-Annual-Report_22Jan24_V1.1.pdf

13. National Health Service. NHS Digital. Accessed 14th February. www.digital.nhs.uk.

14. NHS Digital. The processing cycle and HES data quality. Accessed 14th February 2024. https://digital.nhs.uk/data-and-information/data-tools-and-services/data-services/hospital-episode-statistics/the-processing-cycle-and-hes-data-quality.

15. Kriens O. Lahshal: A concise documentation system for cleft lip, alvolus and palate diagnoses. In: Kriens O, ed. What Is Cleft Lip and Palate? A Multidisciplinary Update Workshop, *Bremen*. Thieme; 1987.

16. ICD10. ICD10 Data. Accessed 14th February 2024. https://www.icd10data.com.

17. Sharp GC, Ho K, Davies A, et al. Distinct DNA methylation profiles in subtypes of orofacial cleft. Clin Epigenetics. 2017;9(1):1–17. doi:10.1186/s13148-017-0362-2

18. Grosen D, Chevrier C, Skytthe A, et al. A cohort study of recurrence patterns among more than 54000 relatives of oral cleft cases in Denmark: Support for the multifactorial threshold model of inheritance. J Med Genet. 2010;47(3):162–168. doi:10.1136/jmg.2009.069385

19. Ludwig KU, Mangold E, Herms S, et al. Genome-wide meta-analyses of nonsyndromic cleft lip with or without cleft palate identify six new risk loci. Nat Genet. 2012;44(9). doi:10.1038/ng.2360

20. Curtis SW, Chang D, Sun MR, et al. FAT4 identified as a potential modifier of orofacial cleft laterality. Genet Epidemiol. 2021;45(7):721–735. doi:10.1002/gepi.22420

21. Greenland S, Senn SJ, Rothman KJ, et al. Statistical tests, P values, confidence intervals, and power: a guide to misinterpretations. Eur J Epidemiol. 2016;31(4). doi:10.1007/s10654-016-0149-3

22. NHS Health Research Authority. What approvals and decisions do I need? https://www.hra.nhs.uk/approvals-amendments/what-approvals-do-i-need/.

23. Gallagher ER, Siebold B, Collett BR, Cox TC, Aziz V, Cunningham ML. Associations between laterality of orofacial clefts and medical and academic outcomes. Am J Med Genet A. 2018;176(2):267–276. doi:10.1002/ajmg.a.38567

24. Farina A, Wyszynski DF, Pezzetti F, et al. Classification of oral clefts by affection site and laterality: a genotype-phenotype correlation study. Orthod Craniofac Res. 2002;5(3):185–191. doi:10.1034/j.1600-0544.2002.02204.x

25. Letra A, Menezes R, Granjeiro JM, Vieira AR. AXIN2 and CDH1 polymorphisms, tooth agenesis, and oral clefts. Birth Defects Res A Clin Mol Teratol. 2009;85(2):169–173. doi:10.1002/bdra.20489

26. Carlson JC, Taub MA, Feingold E, et al. Identifying Genetic Sources of Phenotypic Heterogeneity in Orofacial Clefts by Targeted Sequencing. Birth Defects Res. 2017;109(13):1030–1038. doi:10.1002/bdr2.23605

27. Yin B, Shi JY, Lin YS, Shi B, Jia ZL. SNPs at TP63 gene was specifically associated with right-side cleft lip in Han Chinese population. Oral Dis. 2021;27(3):559–566. doi:10.1111/odi.13566

28. Tao HX, Shi JY, Lin YS, Yin B, Shi B, Jia ZL. Rs9891446 in NTN1 is associated with right-side cleft lip in Han Chinese population. Arch Oral Biol. 2022;141. doi:10.1016/j.archoralbio.2022.105485

29. Fogh-Anderson P. Inheritance of Harelip and Cleft Palate. Busk; 1942.

30. Mastroiacovo P, Maraschini A, Leoncini E, et al. Prevalence at birth of cleft lip with or without cleft palate: Data from the International Perinatal Database of Typical Oral Clefts (IPDTOC). Cleft Palate-Craniofacial Journal. 2011;48(1):66–81. doi:10.1597/09-217

31. Matern O, Sauleau EA, Tschill P, Perrin-Schmitt F, Grollemund B. Left-sided predominance of hypodontia irrespective of cleft sidedness in a French population. Cleft Palate-Craniofacial Journal. 2012;49(3). doi:10.1597/11-025

32. Letra A, Menezes R, Granjeiro JM, Vieira AR. Defining Subphenotypes for Oral Clefts Based on Dental Development.

33. Marazita ML. The evolution of human genetic studies of cleft lip and cleft palate. Annu Rev Genomics Hum Genet. 2012;13:263–283. doi:10.1146/annurev-genom-090711-163729

34. Dixon MJ, Marazita ML, Beaty TH, Murray JC. Cleft lip and palate: Understanding genetic and environmental influences. Nat Rev Genet. 2011;12(3):167–178. doi:10.1038/nrg2933

35. Stoll C, Alembik Y, Dott B, Roth MP. Associated Malformations in Cases with Oral Clefts.; 2000.

36. ONS. 2011 Census: Key Statistics and Quick Statistics for Local Authorities in the United Kingdom.

37. Wang L, Liu Z, Lin H, Ma D, Tao Q, Liu F. Epigenetic regulation of left–right asymmetry by DNA methylation. EMBO J. 2017;36(20):2987–2997. doi:10.15252/embj.201796580

